# Wide-based Foveal Pit: A Predisposition to Idiopathic Epiretinal Membrane

**DOI:** 10.1101/2020.08.13.20174334

**Authors:** I-Hsin Ma, Chung-May Yang, Yi-Ting Hsieh

## Abstract

**Aims:** To report the anatomical characteristics of wide-based foveal pit and its associations with macular diseases.

**Methods:** Wide-based foveal pit was defined as a foveal base width (FBW) larger than the mean value plus one standard deviation of normal population. Eyes with a wide-based foveal pit were retrospectively collected as the study group, and age- and sex-matched subjects with a normal FBW were recruited as the control group. FBW, area of foveal avascular zone (FAZ) and retinal artery trajectory (RAT) were compared between two groups. The characteristics of the fellow eyes in the study group were also described.

**Results:** Fifty two eyes from 52 patients were identified as having a wide-based foveal pit; 43 (82.7%) were female. Both their FBW (474.7 ± 84.6μm) and area of FAZ (0.50 ± 0.11mm^2^) were significantly larger than in the control group (297.6 ± 42.3 μm and 0.29 ± 0.10 mm^2^, respectively; *p* < 0.001 for both), and they also had a wider RAT than the control group (*p* < 0.001). During follow-up, three eyes had developed idiopathic epiretinal membrane. As for their fellow eyes, they either also had a wide-based foveal pit (11 eyes) or had various macular diseases including idiopathic epiretinal membrane (27 eyes), macular hole (5 eyes) and others (16 eyes).

**Conclusions:** Eyes with a wide-based foveal pit also had a large FAZ and a wide RAT, and they might have a predisposition to idiopathic epiretinal membrane formation. Their fellow eyes also had a predisposition to epiretinal membrane and macular hole.

**Synopsis:** Eyes with a wide-based foveal pit had a predisposition to idiopathic epiretinal membrane formation. Their fellow eyes might also have a wide-based foveal pit and had a predisposition to epiretinal membrane and macular hole.

## Introduction

The fovea is a pitted structure in the center of the macula. At the center of the fovea lies the foveola, which is 0.35 mm in size with a depressed center known as the umbo. Histologically, the foveola lacks inner retinal layers, and is also devoid of vasculature. Photoreceptors, specifically cone cells, are densely packed in the foveola.^1^ Due to the depletion of inner retinal layers, light is less scattered during transmission, which in turn gives rise to the high spatial resolution of vision in this area.

The invention of optical coherence tomography (OCT) permitted easier assessment of the structure of the retina and facilitated inspection of the contour of the foveal pit.^2 3^ The different contours of the foveal pit have given rise to a variety of methods to describe the foveal pit contour. All such methods — from measuring the dimensions of the base, slope and shoulder to using mathematic approaches to fit the foveal pit contour into reverse Gaussian equations — have demonstrated the complexity of outlining the foveal pit.^4 5^ Given the many different descriptions, controversy remains to whether we can define certain parameters to differentiate normal foveal contours from pathological ones. Clinically, we have noticed that eyes with a wide-based foveal pit have normal vision without metamorphopsia, but their fellow eyes had either a similar wide-based foveal pit or various kinds of macular diseases. Therefore, we believed such type of foveal contour merited further study.

Some previous study has shown that as compared with the normal fellow eye, patients with macular hole have a widened retinal artery trajectory (RAT), the curve composed of the optic nerve center and the paths of both major temporal retina arteries.^6^ The possible mechanism could be the tangential tractional forces on the vitreomacular interface. We speculated that eyes with a wide foveal base might also be related to an abnormal RAT. In the present study, we collected information on eyes with a wide-based foveal pit as well as their fellow eyes; we described their anatomical characteristics and compared them with normal subjects.

## Material and Methods

### Study Subjects

We retrospectively reviewed consecutive patients who had received macular OCT and OCT angiography (OCTA) using Optovue Avanti RTVue XR OCT (Optovue, Inc., Fremont, CA, USA) in single ophthalmologist’s clinic (Y-T Hsieh) at the National Taiwan University Hospital between November 2016 and August 2018. During the first stage, we chose 50 male and 50 female patients with normal macular structures and foveal contours as representation of normal population. These subjects primarily received OCT for preoperative evaluation for cataract surgery, refractive sugery, or routine check-up during the study period. Those with any of the following characteristics were exluded: (1) irregular or asymmetric foveal contour, (2) retinal tissue loss at the fovea or parafoveal area, (3) any vitreomacular or choroidal diseases noted on OCT or fundus photography, (4) presence of posterior staphyloma, (5) symptoms of metamorphopsia and (6) incomplete data for the axial length, fundus images or B scan and en face angiography images on OCT. We then measured the foveal base width (FBW), which was defined as the distance between two intersections of internal limiting membane and a line parallel to the underlying retinal pigment epithelium layer at 10 μm above the lowest point of the foveal pit in the B-scan OCT image (Figure 1). We used both the horizontal and vertical sections of B-scan OCT for measurement, and recorded the mean value as the FBW. Only one eye was randomly selected from each subject for calculation. Of the 100 normal eyes, the mean FBW was 321.8± 67.8 μm. We then defined a wide-based foveal pit as a FBW larger than the mean value plus one standard deviation of normal population, which was 390 μm.

**Figure 1.**
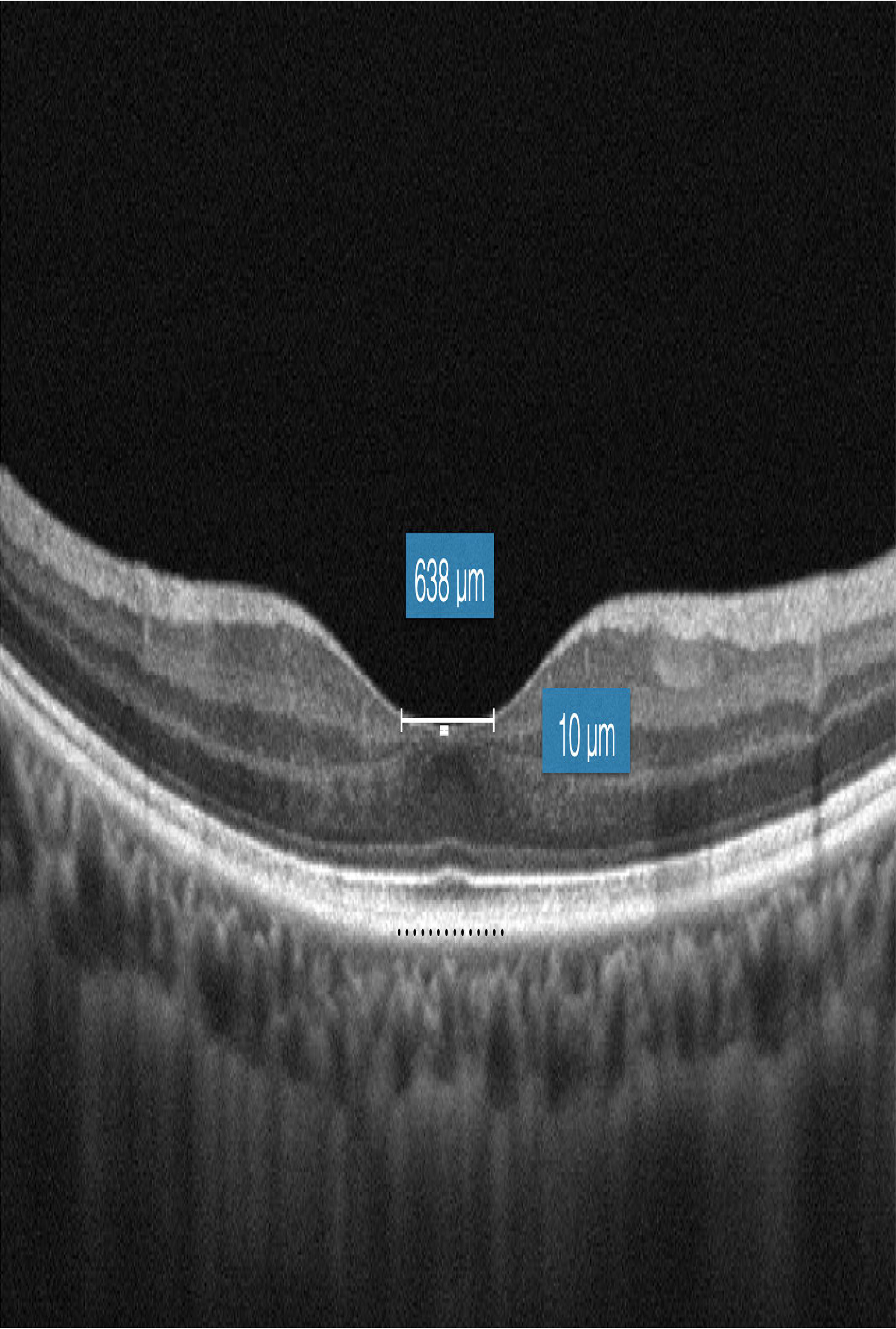
Measurement of the width of the foveal pit. We drew a line parallel to the retinal pigment epithelial layer (dotted black line) at the level of 10 μm above the deepest point of the foveal pit, then measured the distance between its intersections with the retinal inner surface as the width of the foveal pit (638 μm in this example).

During the second stage, we retrosepctive collected eyes with a wide-based foveal pit during the same period. The exclusion criteria were the same as the normal population in the first stage. For subjects whose bilateral eyes both matched the criteria for wide-based foveal pit, only one eye was enrolled in the study group. We also collected information on the fellow eyes of the study group for analysis. As for the control group, we recruited age- and sex-matched subjects (same sex, age difference < 5 years) who had a FBW equal to smaller than 390 μm during the same period. We collected the demographics and clinical characteristics, including age, sex, axial length, status of posterior vitreous detachment and any macular diseases noted on OCT or fundus images. The horizontal and vertical B scan OCT images and the en face angiography images of the macula were obtained for analysis. We obtained the fundus images using either color fundus photography, fundus auto-fluorescence or fundus image obtained from the OCT machine.

This study was approved by the Ethics Committee and Institutional Review Board of National Taiwan University Hospital and adhered to the Declaration of Helsinki.

### Parameters of the Foveal Pit

The parameters of the foveal pit included the FBW and the area of the foveal avascular zone (FAZ). The measurement of FBW has been described above. The FAZ was outlined as the region without vasculature in the en face OCTA image, and the area of the FAZ was calculated using the built-in algorithm of the Optovue Avanti RTVue XR OCT.

### Retinal Artery Trajectory (RAT)

We measured RAT using a modified version of the method proposed by Yoshihara et al.^6^ First, we rotated the fundus images 90 degrees clockwise for right eyes and 90 degrees counterclockwise for left eyes using ImageJ (version 1.47, National Institutes of Health, Bethesda, MD, USA; available at: http://imagej.nih.gov/ij/). We then manually dotted along the arcade arteries, placing the first dot at the site where the retinal artery emanates from the optic disc. We labeled each fundus photograph with 20 to 24 dots and generated the x-y coordination with the “invert Y coordinate” function of ImageJ. We then fitted those coordinates to the best fit curve with a second-degree polynomial equation. The coefficient of the second-degree polynomial represents the width of the artery curve, with a larger coefficient representing a narrower curve (Figure 2). All RAT measurements were performed by the same person (I-Hsin Ma), and repeated measurements of RAT was done in seven eyes at different time to verify the measurement bias.

**Figure 2.**
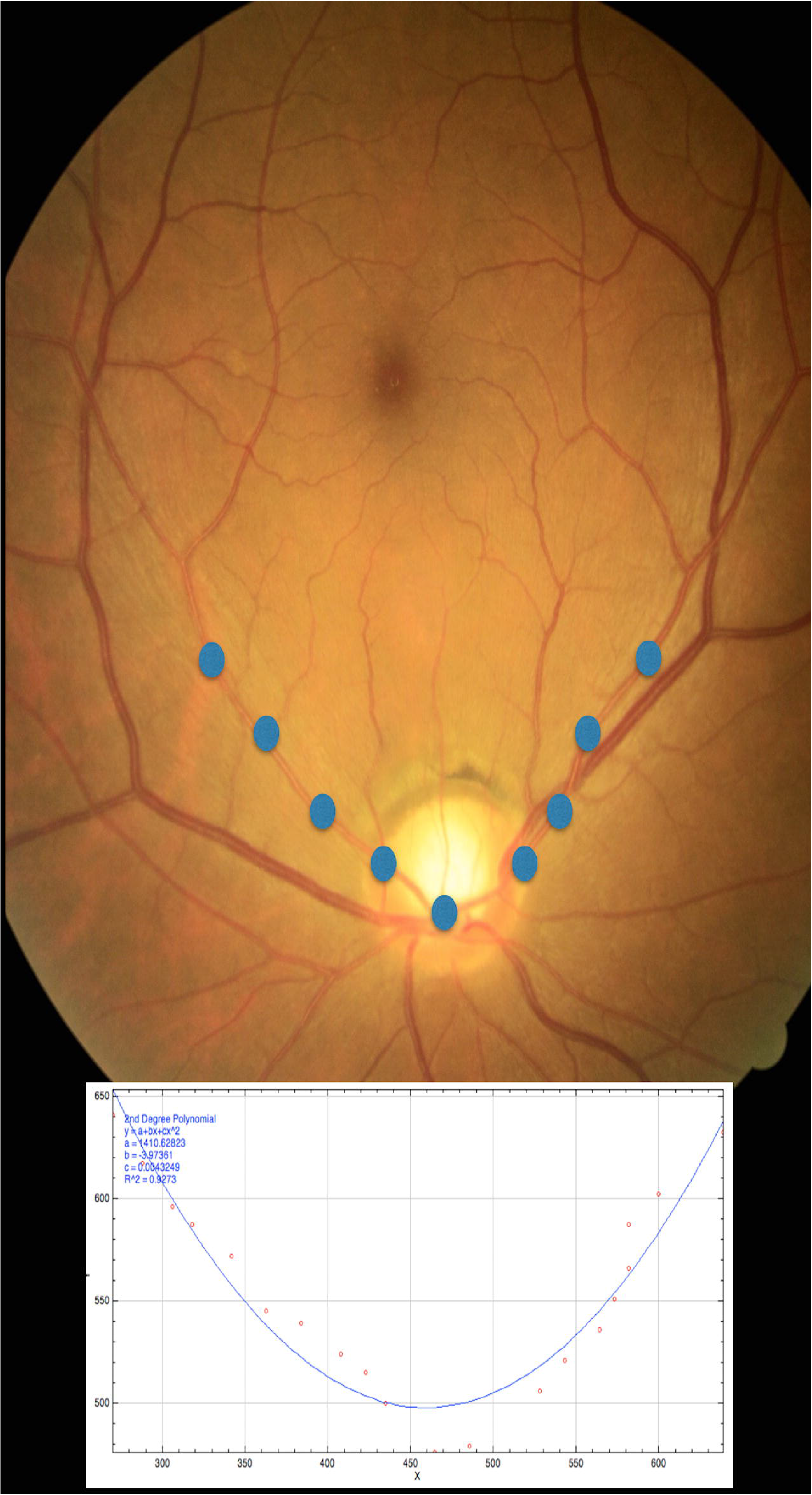
Measurement of the retinal artery trajectory (RAT) from color fundus photography. We manually placed 20 to 24 dots along the arcade arteries, with the first at the site where the retinal artery emanates from the optic disc (sample blue dots shown). These dots then fitted to the best fit curve with a second-degree polynomial equation using the “invert Y coordinate” function of ImageJ (lower part).

### Statistical Analysis

We used paired t-tests to compare continuous variables (axial length, FBW, area of FAZ and RAT) between the study group and their fellow eyes, as well as between the study group and their paired matches in the control group. We also used paired-*t* tests to examine the difference between repeated measurements of RAT. SPSS software (SPSS 22.0; IBM Corp., Armonk, NY, USA) was used for data analysis. A *p* value of less than 0.05 was statistically significant. A Bland-Altman plot was drawn to present the intersessional variation of RAT measurement.

## Results

### Demographics

We enrolled a total of 52 eyes with a wide-based foveal pit from 52 patients in the study group. The mean age was 63.4 ± 9.3 years, and 43 of participants (82.7%) were female. The mean axial length was 23.9 ± 1.85mm and 38 eyes (73%) had complete posterior vitreous detachment. The control group contained 52 eyes in 52 patients who were matched to the study group by age and sex. No differences in age or axial length between the study group and the control group were noted (*p* > 0.05 for both) (Table 1).

**Table 1.** Clinical characteristics of eyes with a wide-based foveal pit (study group) and those in the control group.

|  | Study group<br>(n=52 ) | Control group<br>(n=52) | *P* value |
| --- | --- | --- | --- |
| Age (year) | 63.4± 9.3 | 61.1 ±10.4 | 0.87 |
| Axial length (mm) | 23.9± 1.85 | 23.75±0.93 | 0.38 |
| Foveal base width (μm) | 474.7± 84.6 | 297.6± 42.3 | <0.001 |
| Area of FAZ (mm<sup>2</sup>) | 0.50± 0.11 | 0.29± 0.10 | <0.001 |
| Retinal artery trajectory | 0.18± 0.11 | 0.31± 0.14 | <0.001 |

FAZ: foveal avascular zone.

### Parameters of Foveal Pit and RAT

The mean FBW in the wide-based group was 474.7± 84.6μm, which was significantly larger than that in the control group (297.6 ± 42.3 μm, *p* < 0.001). The mean FAZ area was 0.50± 0.11mm^2^ in those with a wide-based foveal pit, which was also significantly larger than that in the control group (0.29 ± 0.10 mm^2^, *p* < 0.001). As for the RAT, the mean coefficient was 0.18 ± 0.11 in the study group and 0.31 ± 0.14 in the control group (p < 0.001), indicating that those with a wide-based foveal pit had a wider RAT than did the normal controls (Table 1). The intersessional variability of RAT was small as demonstrated in the Bland-Altman plot (Supplementary Figure 1).

### Development of Idiopathic Epiretinal Membrane

Three eyes with a wide-based foveal pit had developed idiopathic ERM at their follow-up examinations (Figure 4). In one case, the ERM was only visible on B-scan OCT with mild disturbance of the foveal pattern; the other two eyes developed ERM, which also narrowed the RAT with a noticeable increase of the coefficient from 0.38 to 0.57 in one and from 1.35 to 1.53 in the other.

**Figure 3.**
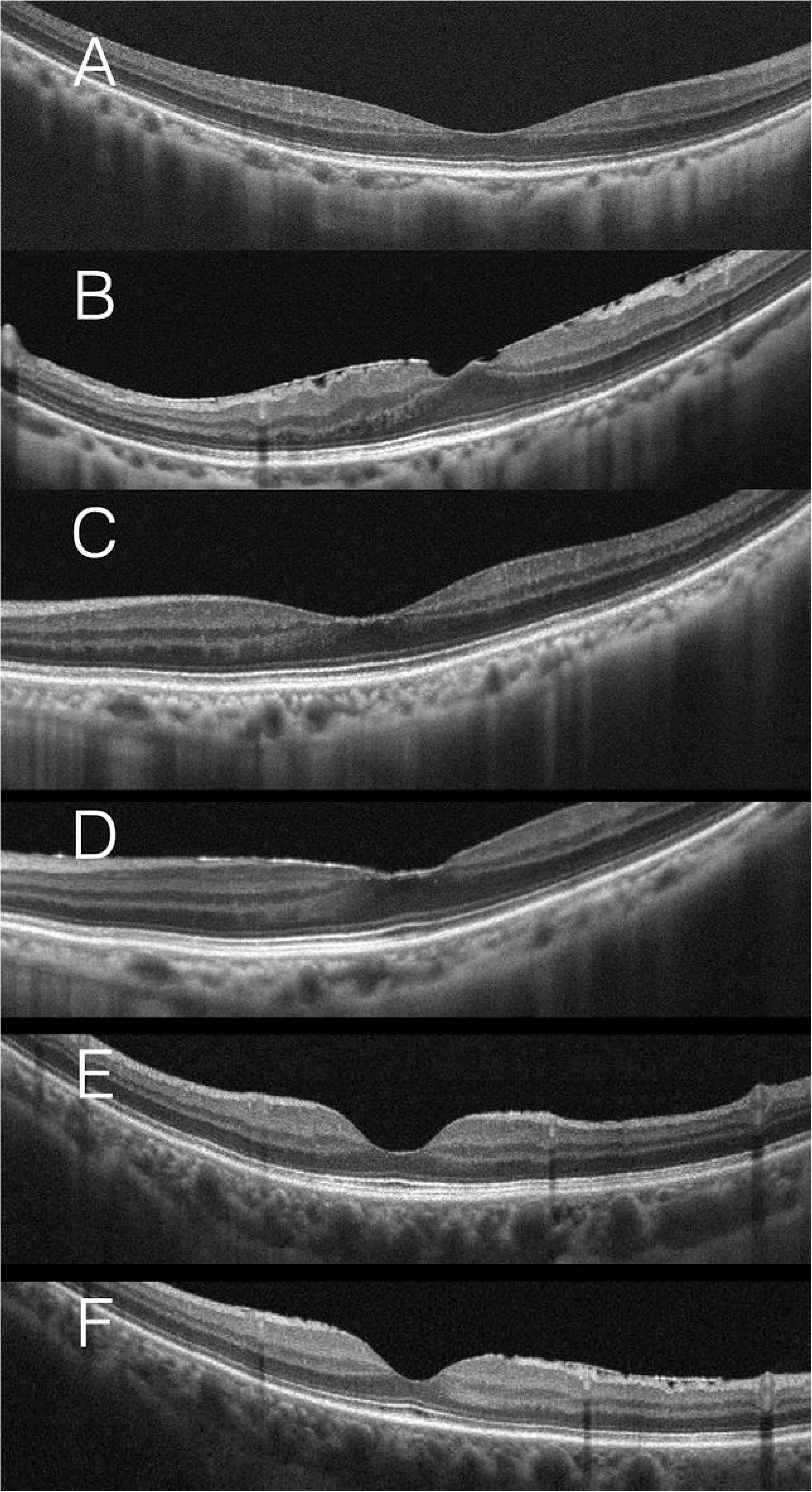
Sequential optical coherent tomography scans of three cases with a wide-based foveal pit showed idiopathic epiretinal membrane formation during the follow-up examinations. A,C, E: at baseline; B: 12 months later of A; D: 12 months later of C; F: 18 months later of E.

### Clinical Characteristics of Fellow Eyes

Of the 52 fellow eyes in the study group, many had various macular diseases, including epiretinal membrane (ERM) (27 eyes), macular hole (MH) (5 eyes), retinal vein occlusion (RVO) (5 eyes), retinal pigment epithelium detachment (4 eyes), choroidal neovascularization (3 eyes), central serous chorioretinopathy (2 eyes), choroidal hemangioma (1 eye) and polypoidal choroidal vasculopathy (1 eye). Eleven fellow eyes also had a wide-based foveal pit, including all fellow eyes without any specific vitreoretinal diseases (4 eyes). For the fellow eyes with ERM, the mean coefficient of RAT was 0.30, similar to that in the control group (0.35). On the other hand, the mean coefficient of RAT was 0.14 in the fellow eyes with MH and 0.21 in those with RVO, similar to that in eyes with a wide-based foveal pit (0.18) (Table 2).

**Table 2.** Area of foveal avascular zone and retinal artery trajectory in different groups. ERM: epiretinal membrane; FAZ; foveal avascular zone; MH: macular hole; RVO: retinal vein occlusion.

|  | Wide-based<br>foveal pit | RVO | MH | ERM | Control<br>group |
| --- | --- | --- | --- | --- | --- |
| Area of FAZ<br>(mm<sup>2</sup>) | 0.50 ± 0.11 | 0.53 ± 0.15 | 0.42 ± 0.18 | 0.18 ± 0.14 | 0.32 ± 0.17 |
| Retinal artery<br>trajectory | 0.18 ± 0.11 | 0.21 ± 0.11 | 0.14 ± 0.03 | 0.30 ± 0.21 | 0.35 ± 0.14 |

## Discussion

In this study, we described a specific type of foveal shape: wide-based foveal pit. To define a wide-based foveal pit, we have enrolled 100 normal subjects to calculate the distribution of FBW in normal population. Then we used the mean value plus one standard deviation as the cut-off point for the lower limit of FBW in a wide-based foeval pit. Because the contour of the foveal pit varies widely in a normal population, clinicians usually treat the wide-based foveal pit as a normal variant, especially as such eyes usually have normal vision without metamorphopsia. However, we found some of them developed idiopathic ERM later, and their fellow eyes either also had a wide-based foveal pit or had some macular diseases including idiopathic ERM, MH and RVO. Thus, we think that both eyes of the same individual might have had similar foveal contours originally, but later some of them evolved into some macular diseases. Rather than a normal variant, we hypothesize that the wide foveal base represents a pathological change that predisposes the eye to other macular diseases.

The reported prevalence of ERM ranges 2-18% in general populations of different regions and ethnicities.^7-12^ Age-stratified results revealed a higher prevalence of ERM formation in the elderly, with a prevalence ranging 15.1-35.7% in those older than 70 years.^8 12^ In our study group, the mean age was 63.2 years while the proportion of ERM in the fellow eyes was 51.9%, a rate much higher than that in the general population. The prevalence of MH in the fellow eyes (9.6%) was also much higher than that in the normal population (0.02-0.8%) according to several large population-based studies.^11 13-15^ Although our study did not include the general population, such results still suggest some correlation between wide-based foveal pit and these diseases. Furthermore, we also found that females predominated in eyes with a wide-based foveal pit (82.7% female vs. 17.3% male), which is an interesting finding because females also reportedly predominate in cases of idiopathic ERM, MH and RVO.^16-18^ This result implies that the macula of females may have characteristics that act as common predisposing factors for wide-based foveal pit and some macular diseases including idiopathic ERM, MH and RVO.

For eyes with a wide-based foveal pit, that had not only a large FAZ but also a wide RAT. This result implies that some centrifugal force may exist in these eyes that results in both a wide foveal abse and a wide RAT. The relationships among ERM, MH, wide RAT and a wide-based foveal pit are intriguing and closely related. Yoshihara et al.^6^ reported an association of MH with a wider RAT. They believe that a tangential force may widen the major retinal vessels and also transmit to the central foveal area. As a consequence, a wide RAT would result in the formation of MH. Our study results support their theory; we found an association of the presence of wide-based foveal pit with a wide RAT, and that their fellow eyes had a predisposition to MH. We propose that the tangential centrifugal force on the vitreomacular interface will exert tangential traction on the retinal arcade vessels and lead to the widening of RAT; it will also exert tangential traction on the central macula, which will widen the foveal base. Over time, the centrifugal force continues to drag the macula and MH may eventually develop.

ERM is composed of glia, fibrocytes, myofibrocytes, retinal pigment epitheliums and sometimes hyalocytes.^19^ Two types of pathogenesis for ERM have been proposed: one states that ERM comes from cellular growth and collagen deposition along the cleft of the internal limiting membrane (ILM); and the other states that ERM develops along the posterior scaffold of the hyaloid membrane, so that ERM can be detected in eyes with either a detached or attached posterior hyaloid.^20-23^ Our study results support the former type of pathogenesis for ERM. We propose that, in eyes with a wide-based foveal pit and a wide RAT, the centrifugal force on the macular surface may also disrupt the ILM over the macula, which then results in some trivial and clinically undetectable ILM deficits. Such deficits may then give rise to glial cell proliferation and ERM formation. After its formation, the contractile characteristic of ERM will result in a centripetal force on the macula, which will narrow the RAT. We observed this result in our finding that three eyes with a wide-based foveal pit developed ERM during the follow-up period and, as we expected, the RAT was narrowed after the formation of ERM. According to the above findings, we further propose that ERM formation may act as a natural protection mechanism against MH formation in eyes with centrifugal traction on the macula. In the absence of ERM formation over the fovea in eyes with a wide-based foveal pit, the centrifugal force may persist and MH may eventually develop. Our study results support this assertion in that the fellow eyes with MH had a similar RAT as the eyes with a wide-based foveal pit, while the fellow eyes with ERM had a much narrower RAT. We will continue to follow the patients with a wide-based foveal pit to monitor for any subsequent MH formation or more ERM formation.

In this study, we used the en face OCTA images to measure the area of FAZ. We measured it through the ILM to the outer plexiform layer, including the superficial, intermediate and deep retinal capillary plexus in normal structures. We found that the mean FAZ was significantly different among eyes with wide-based foveal pit, those with different diseases and those of normal controls. Eyes with ERM had the smallest FAZ, while those with MH or RVO had a larger FAZ, which was similar to the eyes with a wide-based foveal pit. Previous studies have demonstrated that, during ocular development, the formation of FAZ precedes the formation of the foveal pit.^24 25^ The model of foveal pit development suggests that intraocular pressure and retinal stretching during ocular growth will widen the foveal pit. Having less retinal tissue, the FAZ is more susceptible to further stretching than is the vascularized retina, and is therefore more deformable. Thus, a larger FAZ was usually associated with a wider and thinner fovea.^26 27^ These findings further supported our proposal that a large centrifugal stretching force during ocular development will result in RAT widening, FAZ enlargement and the subsequent widening of the foveal pit. As time progresses, MH may then develop; otherwise, glial proliferation and ERM formation may be induced with subsequent narrowing of the RAT and FAZ, which may protect against MH formation.

In addition to ERM and MH, we also found an association of wide-based foveal pit with RVO in the fellow eyes. It has been reported that patients with unilateral RVO had enlarged FAZs in both the RVO eyes and the non-RVO ones, when compared with normal controls.^28^ The authors proposed that the eyes without RVO may also have parafoveal capillary dropout, which resulted in FAZ enlargement. However, our study results indicate a possible association of RVO with widening of the foveal pit and anatomical FAZ enlargement, not capillary dropout in the fellow eye. Further studies are needed to elucidate the underlying mechanism for the association between wide-based foveal pit and RVO.

The major limitation of this study is that this is not a population-based study. We collected cases of the study group as well as the control group from patients who visited our clinic for various reasons, and this selection process may have resulted in selection bias and overestimation of the prevalence of macular diseases in the fellow eyes. However, many of the cases in this study were actually found during routine OCT examination before refractive surgery or cataract surgery. Nevertheless, we do not claim the prevalence, but only a predisposition to the associated diseases. The strength of this study is that the cases in the control group were matched to those in the study group by age and sex, and all cases were of chinese ethnicity. In this way the confounding effect of age, sex and race could be reduced.

In conclusion, we described the anatomical and clinical characteristics of a special type of foveal contour, the wide-based foveal pit. Eyes with a wide-based foveal pit also had a large FAZ and a wide RAT, and female dominated in these subjects. Some eyes with a wide-based foveal pit developed idiopathic ERM at the follow-up examinations. Their fellow eyes also had a predisposition for idiopathic ERM and MH. We propose that the anatomical characteristics of wide-based foveal pit is associated with the formation of idiopathic ERM and MH.

## Data Availability

Data will be available under request.

## Acknowledgement

* Funding statement:

There was no financial or material support for this study.

*Conflict of interest statement:

There were no potential conflicts of interest involving the work under consideration for publication. There were no relevant financial activities outside the submitted work. There were no other relationships or activities that readers could perceive to have influenced, or that give the appearance of potentially influencing what is written in the submitted work.

Contributions and Access to data:

Yi-Ting Hsieh had full access to all the data in the study and takes responsibility for the integrity of the data and the accuracy of the data analysis.

---

**Supplementary Figure 1.**
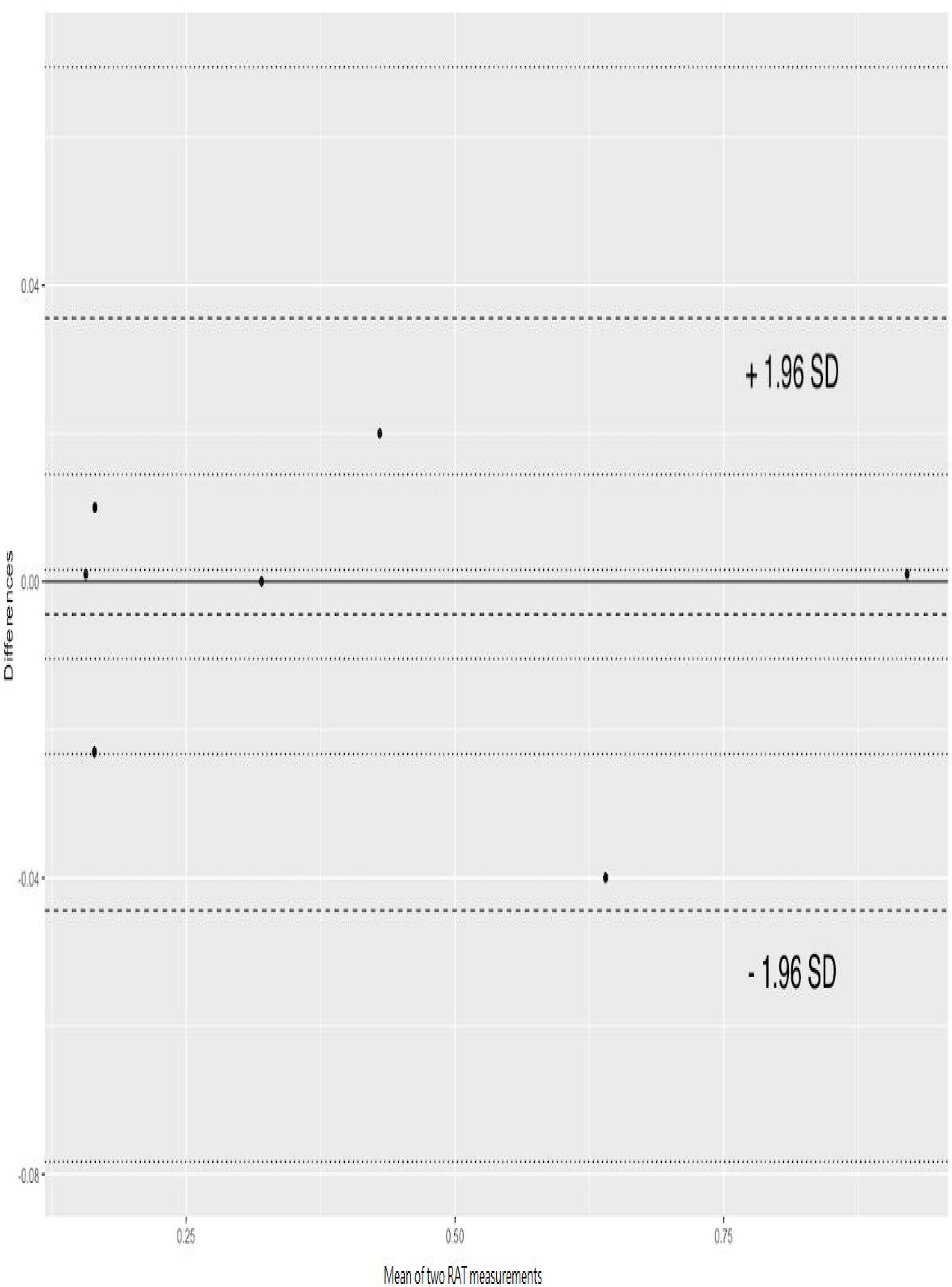
Bland-Altman plot for repeated mesurements of retinal artery trajectory.

